# Sputum scarcity among adolescents and adults with presumptive tuberculosis: a systematic review and meta-analysis

**DOI:** 10.1101/2025.11.02.25339326

**Authors:** Mary Gaeddert, Penelope Papadopoulou, Jennifer Habbes, Tobias Niederegger, Lukas Schik, Julian Meister, Maurizio Grilli, Ashlyn Beecroft, Mikashmi Kohli, Madhukar Pai, Claudia M. Denkinger, Florian M. Marx, Ankur Gupta-Wright

## Abstract

**Background:** The diagnosis of tuberculosis (TB) typically relies on being able to produce a sputum sample for microbiological testing. However, sputum scarcity, the inability to self-expectorate an adequate sputum sample for TB testing, is a well-known concern. Our systematic review and meta-analysis investigated the proportion of sputum scarcity among adolescents and adults being evaluated for presumptive TB in healthcare facilities.

**Methods:** We searched PubMed, Embase, Cochrane Library, Web of Science, and clinical trials databases with no language restrictions from January 2010 to October 2023 using terms for ‘TB’, sputum, and diagnostic studies. We excluded studies with participants aged <15 years, that enrolled patients already providing sputum, or not adequately reporting information on sputum collection. Published summary data was extracted, and the risk of bias was assessed. Summary estimates for the proportion of sputum scarcity were calculated overall and by pre-specified sub-groups. The pooled proportion of sputum scarcity was calculated by random effects meta-analysis. The review protocol was registered on PROSPERO (CRD42023473882).

**Findings:** Our search identified 9895 records, of which 114 studies were included and 81 were rated as a low risk of bias. The median proportion of sputum scarcity across all 114 studies was 6.0% (95% CI: 2.9-9.1%, IQR: 0-19.9%). In subgroup meta-analyses limited to studies collecting one or two self-expectorated spot sputum samples, the pooled estimate of sputum scarcity was 23% (95%CI: 14-33%, n=27). Sputum scarcity was higher in PLHIV sub-groups. The pooled estimate of sputum scarcity in studies enrolling only PLHIV was 24% (95% CI: 15-33%, n=9) for collection of one or two self-expectorated spot sputum samples. Sputum scarcity was the highest in PLHIV inpatients or with advanced disease, with 32% (95% CI: 22-41%, n=5) unable to provide one or two self-expectorated samples. Patients without HIV had the lowest pooled estimate of scarcity, with 12% (95% CI: 3-21%, n=5) unable to provide a self-expectorated sample of any number or collection time. And studies using sputum induction to collect one or two spot samples had a pooled scarcity of 10% (95% CI: 0-21%, n=12).

**Interpretation:** Sputum scarcity is seen in nearly a quarter of patients being evaluated for TB and this compromises TB detection. These findings support the ongoing work to develop non-sputum TB tests.

**Funding:** Gates Foundation (INV-069540)

**Research in Context:** 

**Evidence before this study:** Current diagnostics for tuberculosis (TB) rely on testing sputum samples, and patients unable to produce sputum at the time of evaluation may have a delayed or missed diagnosis. However, there are no reliable estimates about how many patients being evaluted for presumptive TB are unable to produce a sputum sample (sputum scarce) and how this varies in different settings and populations.

**Added value of this study:** This systematic review included data from 114 studies, and found approximately 24% of patients with presumed TB could not produce one or two sputum samples at the time of evaluation. We found higher proportions of sputum scarcity in people living with HIV, and in people living with HIV who were hospital inpatients. We found sputum scarcity was lowest in people without HIV.

**Implications of all the available evidence:** Sputum scarcity can impact nearly a quarter of patients being evaluated for TB and is more common in people living with HIV. These data support the need for non-sputum diagnostics.

## Introduction

Tuberculosis (TB) remains the leading infectious disease cause of death worldwide (1), and while TB can be treated, there is currently a considerable gap between developing TB disease and diagnosis and treatment. An estimated 10.8 million people fell ill with TB in 2023, of whom 2.7 million (25%) were not diagnosed (1). Individuals who do not receive a diagnosis and treatment for TB can remain infectious, contributing to TB transmission, and have considerable morbidity and risk of mortality. Furthermore, globally 40% of TB treatment is started without bacteriological confirmation, and this group appears to have higher risk of poor outcomes (2).

Currently, bacteriological diagnosis of TB typically relies on being able to produce sputum, mucus coughed up from the respiratory tract, which is then tested by smear microscopy, molecular assays or mycobacterial culture (3). Sputum scarcity is the inability to self-expectorate an adequate volume and quality (i.e. sputum rather than saliva) for TB testing (4). There are known challenges with obtaining sputum samples, particularly in children (5) and people living with HIV (PLHIV) (4). In most clinical settings, it is recommended that patients being evaluated for TB provide one or two sputum samples on the spot, i.e. during the same clinical encounter (6). If they are unable to provide sputum at this time, they may return another time for additional attempts, but are at risk of being lost to follow-up (7). In many high TB-burden settings, the use of sputum induction or more invasive pulmonary sampling such as broncho-alveolar lavage is not possible due to availability, cost, infection control risk, and resource limitations, so sputum is primarily collected by self-expectoration (8).

Sputum quality also represents an important aspect of sputum scarcity but determination of what constitutes an adequate sample is difficult. Previous studies indicate that a significant portion of patients provide only saliva-based samples rather than sputum, reducing the sensitivity of diagnostic testing (9, 10). Definitions of sputum quality are heterogeneous (11), and assessment of sputum quality is not regularly reported in diagnostic accuracy studies, potentially underestimating sputum scarcity (12).

Recent target product profiles recognize this need and focus on the development of TB tests using non-sputum samples (13). Research has explored stool (14), urine (15), exhaled breath (16), and oral or tongue swabs (17). While most research shows testing for TB using non-sputum samples often has lower sensitivity than sputum-based tests, non-sputum samples can potentially be collected more readily and from more people, resulting in a higher diagnostic yield and detecting more TB disease overall (18). The frequency of sputum scarcity is not well described, as studies on sputum-based diagnostic tests usually exclude people unable to provide sputum. However, reliable estimates on how many individuals with presumptive TB are unable to produce sputum would enhance understanding of the potential diagnostic yield and impact of non-sputum-based testing for individual and population-based TB control.

The objective of this systematic review was to investigate the frequency of sputum scarcity among adolescents and adults being evaluated for presumptive TB in healthcare facilities, included estimating sputum scarcity in key subgroups, clinical settings, and using different methods of sputum collection. A secondary objective was to investigate the proportion of sputum samples that were not of adequate quality or volume for testing.

## Methods

### Search strategy and selection criteria

Studies were identified through a systematic search of medical databases. The search strategy focused around identifying diagnostic studies conducted in people with presumed TB presenting to healthcare facilities, by combining terms for tuberculosis and diagnostic tests (Appendix Table S1). The search was conducted for all studies from 1 January 2010 to 14 October 2023. In 2010, WHO endorsed the use of Xpert MTB/RIF (Cepheid, USA), a rapid molecular assay for TB diagnosis (19), triggering more intensive research into sputum and non-sputum-based testing for TB. For reasons of feasibility, we did not include studies published before 2010. PubMed, Embase, Cochrane Library, the Web of Science, Literatura Latino Americana en Ciencias de la Salud (LILACS), clinicaltrials.gov, and International Clinical Trials Registry Platform (ICTRP) were searched with no restrictions on language or country. We also identified papers from the references of relevant review articles.

For studies to be included, participants must have attempted to provide samples for TB testing, either as part of study procedures, for example in diagnostic accuracy studies, or via routine TB diagnostic procedures. Studies investigating non-sputum testing were included if sputum-based testing was also conducted. We included randomized clinical trials, cohort studies, and cross-sectional studies, but excluded studies employing active case-finding strategies, prevalence surveys, and household contact studies. Studies enrolling only participants with confirmed TB, extra-pulmonary TB, or those who had already provided sputum prior to screening were excluded. Studies with less than 20 participants overall were also excluded. Studies that enrolled individuals ≥15 years (adolescents and adults) were included, and studies with children <15 years of age were excluded, unless data disaggregated by age were reported.

For title and abstract screening, a sample of 10% were independently screened by at least two reviewers. After reaching concordance between reviewers, only one reviewer independently assessed the remaining titles and abstracts, unless they were unsure in which case a second reviewer was consulted.

### Data extraction

Data was extracted into a piloted and standardized extraction forms. Study quality was assessed using a bespoke tool adapted from the Quality Assessment of Diagnostic Accuracy Studies (QUADAS-2) (13) and JBI tools (20) (Appendix Table S2). Two reviewers (of PP, JH, TN, LS, JM) independently reviewed the full text, extracted data, and completed the quality assessment. Discordance was resolved by decision of a third reviewer (MG). Covidence software (Veritas Health Innovation, Australia) was used for screening, full text review, data extraction and quality assessment. Definitions of study outcomes are given in Table S3.

### Data analysis and synthesis

Characteristics of the included studies were summarized using quantitative data synthesis. Data extracted on the number of participants attempting to provide sputum and the number with a sample successfully collected were used to calculate the proportion of sputum scarcity for each study. When data were reported, the sputum scarcity proportion was adjusted for the number of participants excluded due to providing low volume and/or poor-quality samples as defined by the study. If there was a high degree of heterogeneity between studies as determined by visual assessment of the forest plots and use of *I*^*2*^, an overall summary estimate for the proportion of sputum scarcity was presented as a bootstrapped median with 95% confidence intervals, supplemented by interquartile ranges rather than as a pooled estimate.

To estimate the prevalence of sputum scarcity and to explain likely sources of heterogeneity, meta-analyses were conducted for sub-groups that had at least four studies. The sub-groups specified *a priori* included HIV status, geographic region, clinical setting (e.g. inpatient or outpatient), and level of care (e.g. primary care or referral). We also conducted sub-group analyses of sputum scarcity in PLHIV before initiation of anti-retroviral therapy (ART) as guidelines recommend evaluation for TB before the initiation of ART even in the absence of typical TB symptoms, and studies are often conducted in this group separately from other studies in presumptive TB patients (21). Where sufficient data were available, sub-groups focused on describing the collection of one to two self-expectorated spot sputum samples, as this is the most programmatically relevant.

In a *post-hoc* sensitivity analysis, studies which reported all participants were able to provide a sputum sample were reviewed to assess if there was potential bias towards enrollment of participants who were more likely to provide sputum. We also contacted study authors to clarify sputum collection methods and to ask for additional data where necessary. The median proportion of sputum scarcity was calculated after removing these studies and compared to the result with all studies to estimate how the possible bias may impact the results.

Data analysis was conducted using Stata (StataCorp. 2024. *Stata Statistical Software: Release 17*. College Station, TX: StataCorp LLC.). We used a random effects meta-analysis with the *meta* command package and the random effects restricted maximum likelihood (REML) method to calculate summary estimates and 95% confidence intervals. The review protocol and search strategy were registered on in the PROSPERO database (CRD42023473882) and we followed the PRISMA 2020 guidelines for reporting systematic reviews and meta-analyses (22).

## Results

The combined search of all databases resulted in 9,840 articles after de-duplication. After title and abstract screening, 751 were eligible for full-text review. Of these, 637 were excluded, most commonly for enrolling only participants who had produced sputum or not clearly reporting exclusions for sputum scarcity, leaving 114 studies in the systematic review (Figure 1).

**Figure 1.**
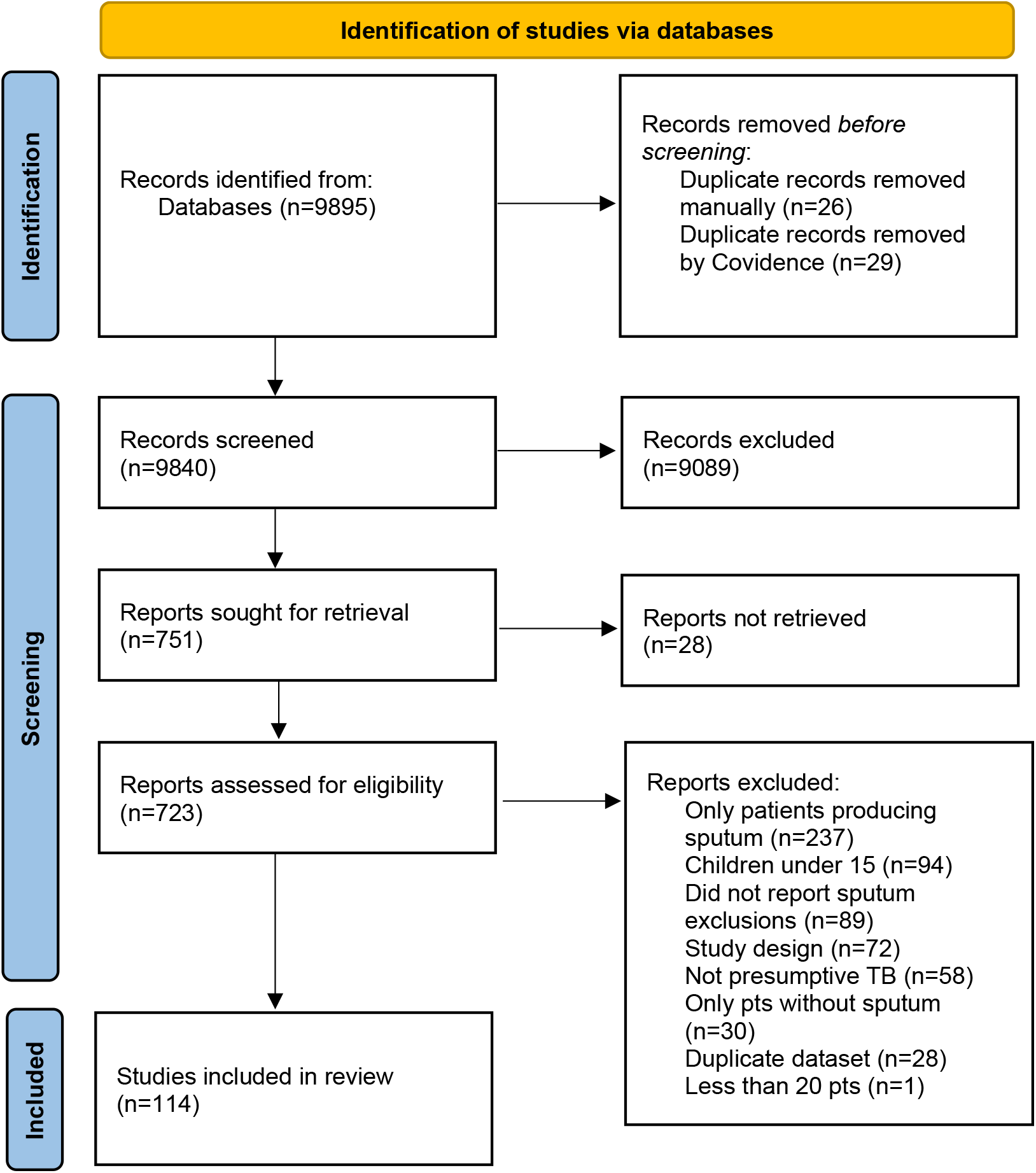
PRISMA flow diagram.

The included studies enrolled a total of 93,158 participants from 37 different countries; 79 (69.3%) studies were conducted in high TB burden countries (23). The majority of studies were cross-sectional in design (n=74, 64.9%) and evaluating the accuracy of TB diagnostic tests (n=65, 57.0%) (Table 1). Most studies enrolled participants from outpatient settings (n=67, 58.8%) at secondary or tertiary level hospitals (n=58, 50.9%). Participants with presumptive TB were enrolled on the basis of presenting symptoms (n=90, 79.0%), being in a high-risk group (n=52, 45.6%), or chest X-ray findings (n=19, 16.7%). Studies enrolling high-risk groups included PLHIV before ART initiation (n=17, 14.9%), PLHIV with advanced HIV disease (n=13, 11.4%), and pregnant women (n=5, 4.4%). Many studies focused enrollment on PLHIV, with 38.6% of studies only enrolling PLHIV (n=44).

**Table 1.**
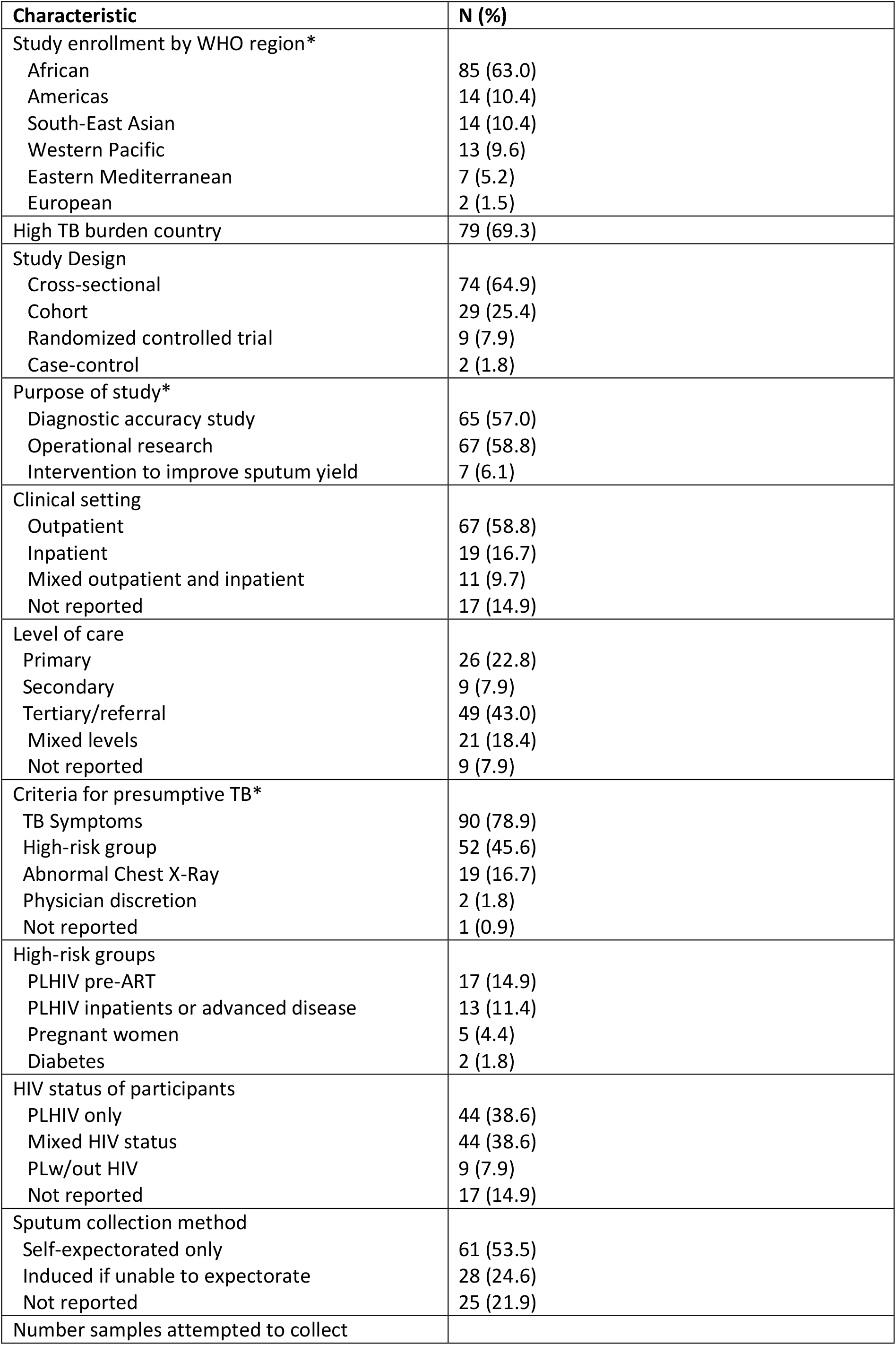
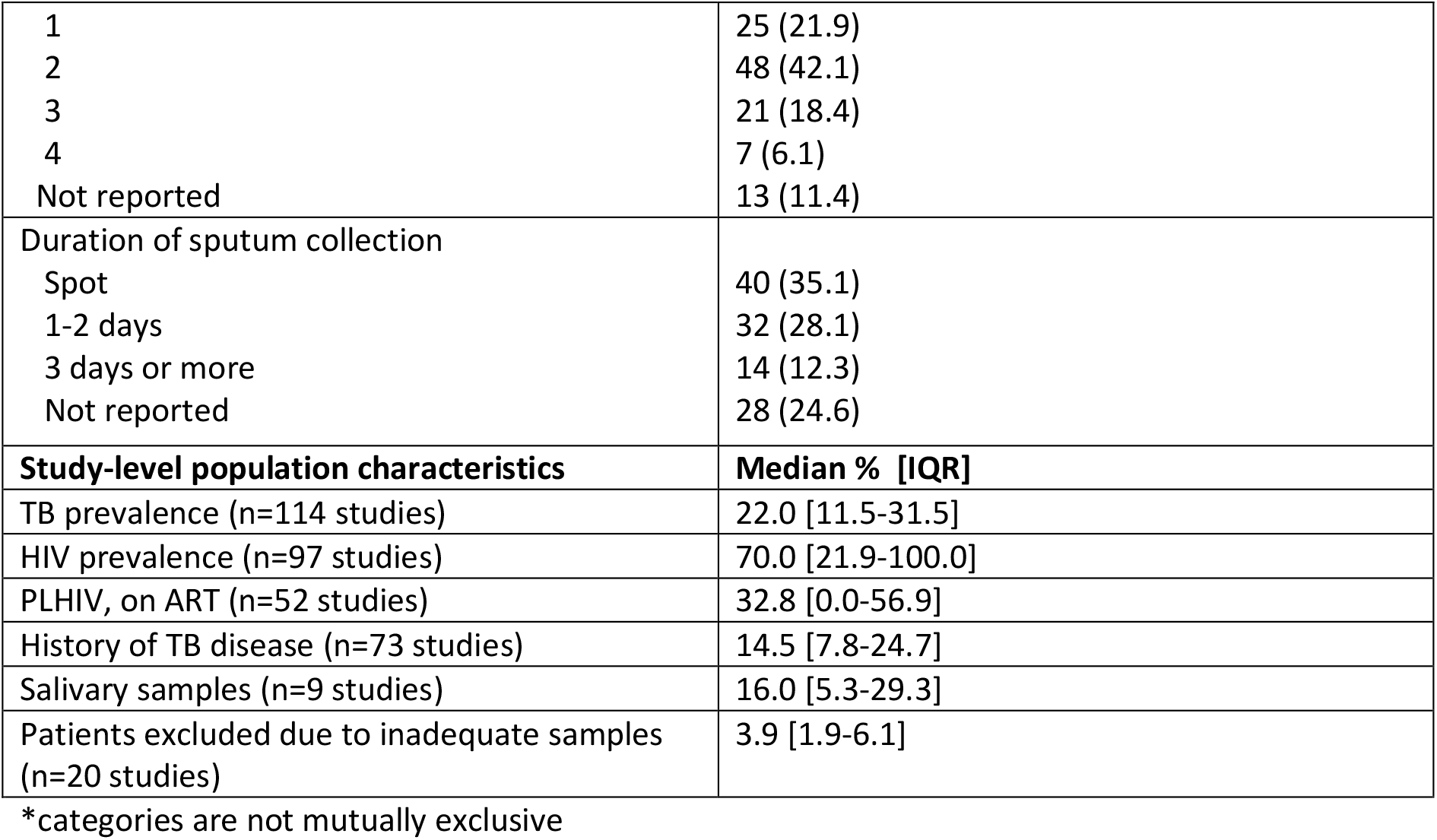
Summary of included studies.

The included studies employed a range of methods for collecting sputum samples. Half of the studies collected only self-expectorated sputum (n=61, 53.5%) or performed sputum induction if participants were unable to self-expectorate (n=28, 24.6%). Most studies attempted to collect one or two sputum samples (n=73, 64.0%) either on the spot (e.g. in one clinical encounter) (n=40, 35.1%) or over the course of one to two days of clinical visits (n=32, 28.1%). However, many studies did not report the method of sputum collection (n=25, 21.9%), the number of samples collected (n=13, 11.4%), or the time of collection (n=28, 24.6%).

The risk of bias assessment scored 109 (95.6%) studies as low risk of bias on the patient selection domain (Figure 2). These studies enrolled a consecutive or random sample of participants, did not use a case-control design, and avoided inappropriate exclusions. For the applicability domain, 81 (71.1%) scored as a low risk of bias. The 31 (27.2%) of studies that scored as a high risk of bias were due to not reporting adequate detail of how sputum samples were collected.

**Figure 2.**
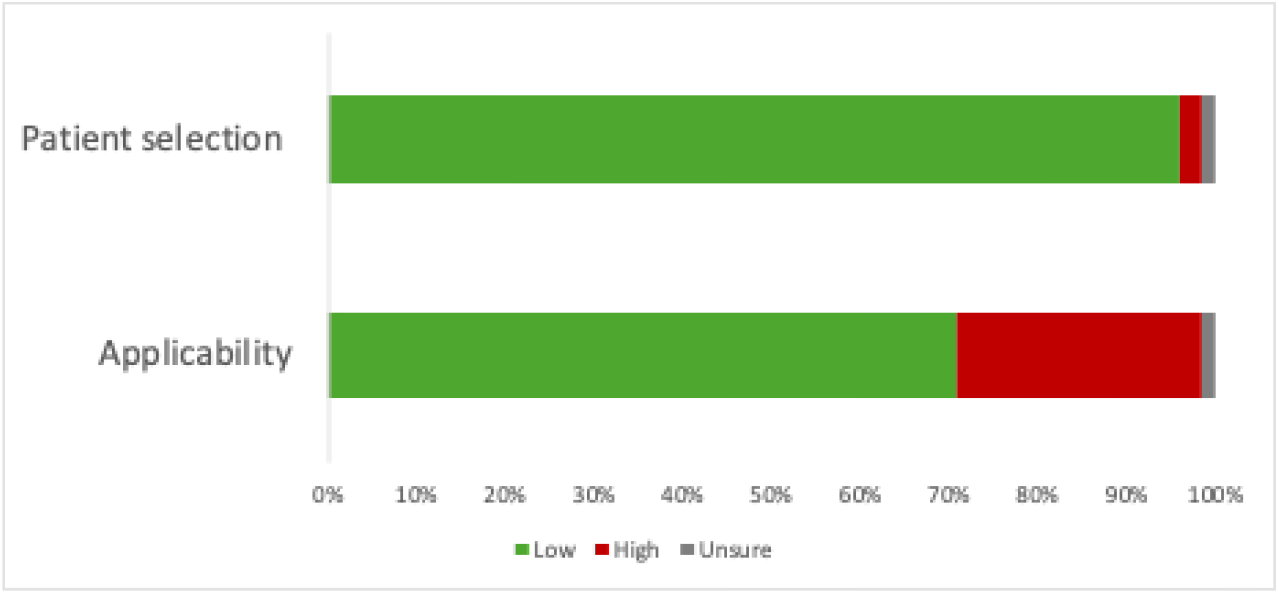
Summary of quality assessment.

Due to the substantial heterogeneity between studies (*I*^*2*^ = 100%), meta-analyses were only conducted for sub-groups. The median proportion of sputum scarcity across all 114 studies was 6.0% (95% CI: 2.9-9.1%, IQR: 0-19.9%). When separated by collection method, the median was 11.4% (95% CI: 4.3-18.5%, IQR 3.3-27.3, n=72) for all studies collecting self-expectorated sputum, and 5.5% (95% CI: 0.3-10.7%, IQR 0.0-10.8, n=28) for all studies using sputum induction if the participant was unable to self-expectorate.

When limited to studies reporting collection of one or two self-expectorated spot sputum samples, pooled sputum scarcity was 23% (95%CI: 14-33%, n=27) (Figure S1, Figure 3). Sputum scarcity was higher in sub-groups with PLHIV compared to those not living with HIV. In studies with results disaggregated by HIV status, the pooled estimate of sputum scarcity in PLHIV was 24% (95% CI: 15-33%, n=9) for collection of one or two self-expectorated spot sputum samples (Figure S2). In populations with mixed HIV status, sputum scarcity was 24% (95% CI: 7-40%, n=11) for collection of one or two self-expectorated spot samples (Figure S3). The median proportion of PLHIV in these studies was 36.7% (IQR: 17.0-66.0%). For PLHIV being evaluated before the initiation of ART, sputum scarcity was 20% (95%CI: 6-33%, n=5) for collection of one or two self-expectorated samples either on the spot or over two days (Figure S4). Patients without HIV had the lowest pooled estimate of scarcity, with 12% (95% CI: 3-21%, n=5) unable to provide a self-expectorated sample of any number or collection time. When analysed by setting, scarcity was the highest in PLHIV inpatients or with advanced disease, with 32% (95% CI: 22-41%, n=5) unable to provide one or two self-expectorated sputum samples over any time (Figure S6). In outpatient settings, 22% (95%CI: 11-33%, n=22) were unable to provide one or two self-expectorated spot sputum samples (Figure S7).

**Figure 3.**
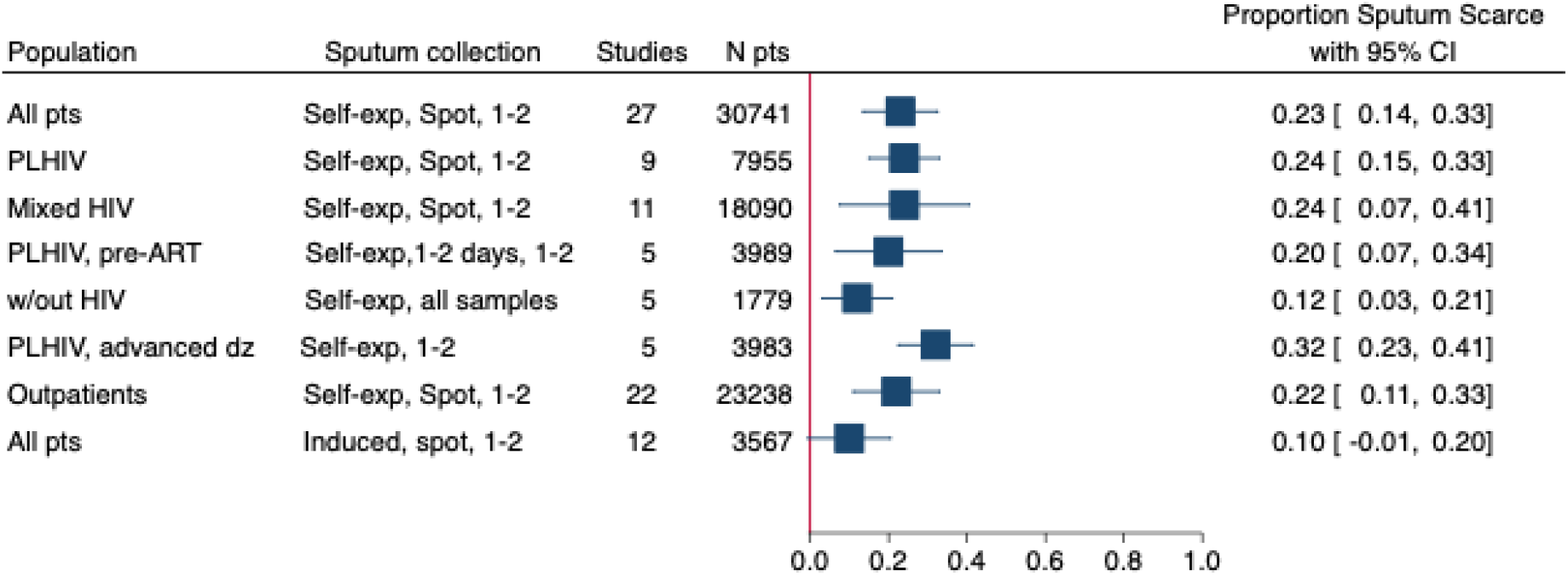
Meta forest plot showing proportion of sputum scarcity for various populations. Legend: sputum collection= method, time, number of samples; N pts = total number of participants included in each analysis

Sputum scarcity was lower when sputum induction was used. Only 10% (95% CI: 0-21%, n=12) were not able to provide one or two induced samples on the spot (Figure S8). Meta-analysis results of additional sub-groups are presented in Table 2. The pooled estimate of sputum scarcity for the collection of self-expectorated sputum in pregnant women was 33% (95%CI: 0-66%, n=5). The pooled estimate of scarcity for collection of one or two self-expectorated spot samples was 22% (95% CI: 10-33%, n=20) in high TB burden countries, compared to 29% (95%CI: 13-44%, n=7) in non-high TB burden countries (Figures S9 and S10).

**Table 2.** Meta-analysis results for additional sub-groups.

| **Population** | **Sputum collection** | **Number of studies** | **Number of participants** | **Proportion Sputum Scarce (95%CI)** |
| --- | --- | --- | --- | --- |
| PLHIV | Induced, Spot, 1-2 samples | 10 | 3,309 | 0.12 (0.0, 0.24) |
| PLHIV pre-ART | Induced, Spot, 1-2 samples | 5 | 1,664 | 0.07 (0.02, 0.13) |
| PLHIV advanced disease | Induced, All samples | 5 | 3,836 | 0.22 (0.01, 0.43) |
| Outpatients | Induced, Spot, 1-2 samples | 7 | 1,829 | 0.07 (0.02, 0.11) |
| Pregnant women | Self-expectorated, all samples | 5 | 5,274 | 0.33 (0.00, 0.66) |
| African region | Self-expectorated, spot, 1-2 samples | 19 | 25,495 | 0.22 (0.12, 0.33) |
| Region of the Americas | Self-expectorated, 1-2 days, 1-2 samples | 5 | 1,079 | 0.15 (0.06, 0.24) |
| Western Pacific region | Self-expectorated, all samples, 1-2 days | 4 | 1,350 | 0.16 (0.0, 0.40) |
| High TB-burden countries | Self-expectorated, spot, 1-2 samples | 20 | 26,285 | 0.22 (0.10, 0.33) |
| Non high TB-burden countries | Self-expectorated, spot, 1-2 samples | 7 | 4,456 | 0.29 (0.13, 0.44) |

Only nine (7.9%) studies reported on the quality of the sputum samples collected, and a median 16.0% (IQR: 5.3-29.3%) of samples were salivary. Of the 20 (17.5%) studies which reported the number of patients providing samples defined as low volume and/or poor-quality, a median 3.9% (IQR: 2.5-6.1%) of patients were excluded due to inadequate samples. The definitions of sample volume and quality that were considered adequate for testing varied by study. Studies using only the Xpert MTB/Rif assay typically excluded samples if less than 1mL in volume, whereas studies performing multiple tests including culture required a higher volume and excluded samples less than 2-4mL.

For studies that reported all participants were able to provide sputum samples, we investigated potential sources of bias in the study design and performed a sensitivity analysis. Of the 18 studies, nine used sputum induction. When removing these studies, the pooled estimate of sputum scarcity for collection of one or two self-expectorated spot samples increases from 24% to 28% (95% CI: 17-39%, n=20) (Figure S11). Additional sensitivity analyses excluding studies with a high risk of bias decreased the pooled estimate to 22% (95%CI: 12-32%, n=21) (Table S5).

## Discussion

This is the first systematic review and meta-analysis to evaluate the ability of adolescents and adults to produce sputum for TB testing. Our main findings are that 24% of people being evaluated for presumed TB are unable to produce one or two spot sputum samples at the time of evaluation. In PLHIV, this was even higher, up to 31% in those with advanced disease or admitted to hospital were unable to provide self-expectorated sputum. Even when sputum induction is performed, 9% of patients still cannot produce a sample at the time of evaluation. Sensitivity analyses investigated potential sources of bias, and when studies reporting everyone was able to provide sputum samples were removed, an estimated 29% of people are unable to produce one or two sputum samples at the time of evaluation.

There was variability in sputum scarcity by population and setting. Scarcity was consistently higher across PLHIV subgroups, especially in PLHIV with advanced HIV disease. This is possibly due to patients being more unwell and therefore physically weak to cough with sufficient force to produce sputum, and/or an impaired inflammatory response in the lungs not promoting sputum production due to the HIV’s impact on the host immune system (24). Correspondingly, the estimates of sputum scarcity were lower people without HIV and being evaluated in outpatient settings, where they presented with presumptive TB. The use of sputum induction allowed more patients to produce sputum, but in subgroup analyses scarcity still ranged from 5-27% across studies using sputum induction.

Sputum quality was reported in only 8% of included studies and a median of 4% of patients were excluded due to provision of inadequate samples. If these findings were consistent across all studies, our estimate of sputum scarcity would likely be higher. There may also be variation in sample quality by patient population but there is insufficient data to address this question. While salivary samples can be tested on molecular platforms such as Xpert MTB/Rif Ultra, previous studies have shown a reduced diagnostic sensitivity (25, 26).

This review provides evidence supporting the need for non-sputum diagnostics and research and implementation of novel samples for TB diagnosis should continue to be advanced. The inability of patients to produce sputum is often not included in studies modelling the cost-effectiveness or impact of new non-sputum diagnostics due to the lack of accurate estimates. The lack of sputum for microbiologic confirmation of TB disease contributes to missed diagnoses, a reliance on empiric TB treatment, and missed opportunities for drug-resistance testing. Introduction on non-sputum sample types, such as tongue swabs, may require different approaches to testing depending on the setting and population (27).

Strengths of this review were the comprehensive literature search and screening methods and the large number of papers included, allowing for broad range of sub-group analyses. This review included many studies from TB high-burden countries, and there was substantial data for PLHIV sub-groups. However, there were less data for populations without HIV and outpatient settings. The wide range of included studies also introduced multiple sources of heterogeneity in the study population, design, and setting, making it difficult to compare studies directly.

This review also included many diagnostic accuracy studies (58%) because they were likely to collect sputum from presumed TB patients. However, these studies may have a bias in screening and enrolling patients who are able to produce sputum, potentially under-estimating the magnitude of sputum scarcity in this population. However, this was balanced by two studies conducted in outpatient settings which reported very high sputum scarcity. We found that the details of sputum collection were often not clearly reported, limiting the data available for the main outcome of one to two self-expectorated spot samples. Studies collecting sputum should consistently report the methods and results of sputum collection, including sample quality. A previous systematic review noted that limited data were reported on sputum quality and definitions were heterogeneous across studies (11).

In conclusion, this review to quantifies the issue of sputum scarcity in adolescents and adults with presumed TB. These findings provide valuable insight into groups that will benefit the most from non-sputum tests that are currently being validated and likely to be reviewed by WHO for policy endorsement.

## Data Availability

All data produced in the present work are contained in the manuscript.

## Declarations

### Contributors

AGW, FM, MP, and CMD developed the idea and initiated the article. MGaeddert, FM, and AGW drafted study design and protocol with input from CMD. The search was designed and conducted by MGrilli, MGaeddert, FM, and AGW. Screening, data extraction and risk of bias assessment was done by MG, PP, JH, TN, LS, and JM. MGaeddert conducted the data analysis. AGW, FM, MGaeddert, AB, MK, MP, and CMD interpreted results. MGaeddert and AGW wrote the first draft of the manuscript. All authors contributed to the manuscript by edits and providing critical feedback. All authors had full access to the data and had final responsibility for submission.

### Data sharing

This manuscript is based on secondary data which is published and publicly available. Data extracted for the analysis is presented in the results section and Supplementary Appendix. The code used for the analysis is available upon request.

## Acknowledgements

This work was supported, in whole or in part, by the Gates Foundation [INV-069540]. The conclusions and opinions expressed in this work are those of the author(s) alone and shall not be attributed to the Foundation. Under the grant conditions of the Foundation, a Creative Commons Attribution 4.0 License has already been assigned to the Author Accepted Manuscript version that might arise from this submission. Please note works submitted as a preprint have not undergone a peer review process.

CMD and AGW are funded by the R2D2 TB Network (National Institute of Allergy and Infectious Diseases of the US National Institutes of Health under award number U01AI152087). AGW is supported by the UK National Institute for Health and Care Research (NIHR305136).

