## Supplemental materials for "Sputum scarcity among adolescents and adults with presumptive tuberculosis: a systematic review and meta-analysis"

**Supplementary Appendix**

### Table S1. Search Strategy

| **PubMed** | |
|---|---|
| **P** | |
| 1 | "Mycobacterium tuberculosis"[MeSH Terms] OR "Tuberculosis"[MeSH Terms] OR "tuberculo*"[Title/Abstract] OR "TB"[Title/Abstract] |
| **I** | |
| 2 | "Sputum"[MeSH Terms] OR "sputum*"[Title/Abstract] |
| **C** | |
| 3 | "Urine"[MeSH Terms] OR "Feces"[MeSH Terms] OR "Blood"[MeSH Terms] OR "Serum"[MeSH Terms] OR "urin*"[Title/Abstract] OR "stool"[Title/Abstract] OR "fece*"[Title/Abstract] OR "Blood"[Title/Abstract] OR "breath*"[Title/Abstract] OR "serum*"[Title/Abstract] |
| **X** | |
| 4 | "Lipoarabinomannan"[Supplementary Concept] OR "Nucleic Acid Amplification Techniques"[MeSH Terms] OR "Lipoarabinomannan"[Title/Abstract] OR "LAM"[Title/Abstract] OR "AlereLAM"[Title/Abstract] OR "FujiLAM"[Title/Abstract] OR "LFLAM"[Title/Abstract] OR "TBLAM"[Title/Abstract] OR "Nucleic Acid Amplification"[Title/Abstract] OR "GeneXpert"[Title/Abstract] OR "xpert*"[Title/Abstract] OR "lamp loop"[Title/Abstract] OR "Truenat MTB"[Title/Abstract] OR "Culture"[Title/Abstract] |
| **Strings**<br>1-4 as in the tables above | |
| 5 | #2 OR #3 |
| 6 | #1 AND #5 AND #4 |
| 7 | #6 AND ("2010/01/01"[PDAT] : "3000/12/31"[PDAT]) |

| **Embase** | |
|---|---|
| **P** | |
| 1 | 'Mycobacterium tuberculosis'/exp OR 'tuberculosis'/exp OR Tuberculo*:ti,ab,kw OR TB:ti,ab,kw |
| **I** | |
| 2 | 'sputum'/exp OR 'sputum examination'/exp OR sputum*:ti,ab,kw |
| **C** | |
| 3 | 'urine'/exp OR 'feces'/exp OR 'blood'/exp OR 'serum'/exp OR 'feces analysis'/exp OR 'urinalysis'/exp OR urin*:ti,ab,kw OR stool:ti,ab,kw OR fece*:ti,ab,kw OR blood:ti,ab,kw OR breath*:ti,ab,kw OR serum*:ti,ab,kw |
| **X** | |
| 4 | 'lipoarabinomannan'/exp OR 'nucleic acid amplification techniques'/exp OR Lipoarabinomannan:ti,ab,kw OR LAM:ti,ab,kw OR AlereLAM:ti,ab,kw OR FujiLAM:ti,ab,kw OR LFLAM:ti,ab,kw OR TBLAM:ti,ab,kw OR "Nucleic Acid Amplification":ti,ab,kw OR GeneXpert:ti,ab,kw OR Xpert*:ti,ab,kw OR "lamp loop":ti,ab,kw OR "Truenat MTB":ti,ab,kw OR Culture:ti,ab,kw |
| Strings | |

| | |
|---|---|
| 1-4 as in tables above | |
| 5 | #2 or #3 |
| 6 | #1 AND #5 AND #4 |
| Publication year from 2010 | |
| 7 | #6 AND [2010-2023]/py |

| | |
|---|---|
| **Cochrane Library** | |
| **P** | |
| 1 | [mh "Mycobacterium tuberculosis"] OR [mh "Tuberculosis"] OR Tuberculo*:ti,ab,kw OR TB:ti,ab,kw |
| **I** | |
| 2 | [mh "Sputum"] OR sputum*:ti,ab,kw |
| **C** | |
| 3 | [mh "Urine"] OR [mh "Feces"] OR [mh "Blood"] OR [mh "Serum"] OR urin*:ti,ab,kw OR stool:ti,ab,kw OR fece*:ti,ab,kw OR blood:ti,ab,kw OR breath*:ti,ab,kw OR serum*:ti,ab,kw |
| **X** | |
| 4 | [mh "Nucleic Acid Amplification Techniques"] OR Lipoarabinomannan:ti,ab,kw OR LAM:ti,ab,kw OR AlereLAM:ti,ab,kw OR FujiLAM:ti,ab,kw OR LFLAM:ti,ab,kw OR TBLAM:ti,ab,kw OR "Nucleic Acid Amplification":ti,ab,kw OR GeneXpert:ti,ab,kw OR Xpert*:ti,ab,kw OR "lamp loop":ti,ab,kw OR "Truenat MTB":ti,ab,kw OR Culture:ti,ab,kw |
| Strings<br>1-4 as in the tables above | |
| 5 | #2 OR #3 |
| 6 | #1 AND #5 AND #4 |
| Publication year from | |
| 7 | publication date from Jan 2010 to present<br>for CENTRAL applied in Endnote (729 > 684) |

| | |
|---|---|
| **Literatura Latino Americana en Ciencias de la Salud (LiLACS)** | |
| **P** | |
| 1 | Tuberculo* OR TB |
| **I / C** | |
| 2 | sputum* OR |
| 3 | urin* OR stool OR fece* OR blood OR breath* OR serum* |
| **X** | |
| 4 | Lipoarabinomannan OR LAM OR AlereLAM OR FujiLAM OR LFLAM OR TBLAM OR "Nucleic Acid Amplification" OR GeneXpert OR Xpert* OR "lamp loop" OR "Truenat MTB" OR Culture |
| Strings<br>1-4 as in the tables above in the fields Title, Abstract, Subject | |
| 5 | 1 AND (2 OR 3) AND 4 |
| | (tuberculo* OR tb) AND (sputum* OR urin* OR stool OR fece* OR blood OR breath* OR serum*) AND (lipoarabinomannan OR lam OR alerelam OR fujilam OR lflam OR tblam OR "Nucleic Acid Amplification" OR genexpert OR xpert* OR "lamp loop" OR "Truenat MTB" OR culture) AND ( db:("WPRIM" OR "LILACS" OR "IBECS" OR "SES-SP" OR "AIM" OR "BINACIS" OR "LIPECS" OR "WHOLIS" OR "CUMED" OR "BDENF" OR "MedCarib" OR "HANSENIASE" OR "BIGG" OR "VETINDEX" OR "BBO")) AND (year_cluster:[2010 TO 2023]) |
| Publication year from 2010 | |
| 6 | Publication date from Jan 2010 to present |

| **Web of Science Core Collection** | |
|---|---|
| **P** | |
| 1 | Tuberculo* OR TB |
| **I** | |
| 2 | sputum* |
| **C** | |
| 3 | urin* OR stool OR fece* OR blood OR breath* OR serum* |
| **X** | |
| 4 | Lipoarabinomannan OR LAM OR AlereLAM OR FujiLAM OR LFLAM OR TBLAM OR "Nucleic Acid Amplification" OR GeneXpert OR Xpert* OR "lamp loop" OR "Truenat MTB" OR Culture |
| Strings<br>1-4 as in the tables above | |
| 5 | #2 OR #3 |
| 6 | #1 AND #5 AND #4 |
| Publication year from | |
| 7 | Publication year from 2010 |

| **ClinicalTrial.gov** | |
|---|---|
| **P** | |
| 1 | tuberculosis OR TB |
| **I / C** | |
| 2 | Sputum OR |
| 3 | urine OR stool OR feces OR blood OR breath OR serum |
| **X** | |
| 4 | Lipoarabinomannan OR LAM OR AlereLAM OR FujiLAM OR LFLAM OR TBLAM OR "Nucleic Acid Amplification" OR GeneXpert OR Xpert OR "lamp loop" OR "Truenat MTB" OR Culture |
| Strings<br>1-4 as in the tables above | |
| 5 | 1 AND (2 OR 3) AND 3 |
| 6 | From 2010 (in Endnote applied) |

| **International Clinical Trials Registry Platform ICTRP (WHO Trials)** | |
|---|---|
| **P** | |
| 1 | tuberculosis OR TB |
| **I / C** | |
| 2 | sputum OR |
| 3 | urine OR stool OR feces OR blood OR breath OR serum |
| **X** | |
| 4 | Lipoarabinomannan OR LAM OR AlereLAM OR FujiLAM OR LFLAM OR TBLAM OR Nucleic Acid Amplification OR GeneXpert OR Xpert OR lamp loop OR Truenat MTB OR Culture |
| Strings (in advanced mode) | |
| 5 | 1 AND (2 OR 3) AND 4 |

**Table S2. Template for Risk of Bias assessment**

| **Domain**<br>**Signaling Question** | **Accepted values and answers** |
|---|---|
| **Domain 1: Patient Selection**<br>Could the selection of patients have introduced bias? | |
| 1. Was a consecutive or random sample of patients enrolled? | Random → yes<br>Consecutive → yes<br>Convenience → no<br>NR → unsure |
| 1. Was a case-control design avoided? | Cohort → yes<br>Cross-sectional → yes<br>Case-control → no<br>NR → unsure |
| 2. Did the study avoid inappropriate exclusions? (e.g. previous TB, pregnancy, HIV) | Yes → yes<br>No → no<br>NR → unsure |
| Scoring:<br>Yes on ≥ 2 questions → Low<br>No on ≥ 2 questions → High<br>Unsure on ≥ 2 questions → Unsure | |
| **Domain 2: Applicability**<br>Is there a concern that the included participants do not match the review question? | |
| 1. Were the study participants and setting described in detail, including information on screening and exclusions? | Yes → yes<br>No → no<br>NR → unsure |
| 2. Were methods of sputum collection and exclusions described in detail? | Yes → yes<br>No → no<br>NR → unsure |
| 3. Were results of sputum collection described in a standard, reliable way for all participants? | Yes → yes<br>No → no<br>NR → unsure |
| Scoring:<br>Yes on ≥ 2 questions → Low<br>No on ≥ 2 questions → High<br>Unsure on ≥ 2 questions → Unsure | |

**Table S3. Study definitions and outcomes**

| Term | Definition | Source |
|---|---|---|
| Self-expectorated sputum | Mucus coughed up from the respiratory tract by the patient without assistance | (1) |
| Sputum induction | Mucus coughed up from the respiratory tract after administration of nebulized saline | (2) |
| Sputum scarcity | The inability to provide an adequate sputum sample for TB testing | (3) |
| Spot samples | Sputum sample provided by self-expectoration during one clinical encounter or visit | (4) |
| Adequate samples | Samples determined to be of sufficient quality and volume required for testing. Definitions vary by study. | |
| Sputum quality | Based on the macroscopic appearance of the sample. Standard categories are salivary, mucoid, purulent, and blood-stained. | (5) |
| Sputum volume | Minium volume needed for diagnostic testing. May vary based on tests used. | (6, 7) |

### Table S4. Characteristics of included studies

| Study ID | Design; DTA | Country (-ies) | Clinical setting | Healthcare level | N pts | TB prevalence (%) | HIV prevalence (%) | Previous TB (%) | Collection method | Number samples | Collection time | Sputum scarce, 95% (%) | Salivary (%) | RoB |
|---|---|---|---|---|---|---|---|---|---|---|---|---|---|---|
| Acharya, 2022 (8) | CSS; Yes | India | Mixed | Mixed | 201 | 21.0 | 100 | NR | NR | NR | NR | 0.36 (0.30, 0.43) | NR | Low, Low |
| Adelman, 2015 (9) | CSS; no | Ethiopia | Outpatient | Tertiary | 256 | 6.0 | 100 | 33.0 | Self-exp | 3 | 2 days | 0.15 (0.11, 0.20) | NR | Low, Low |
| Ahsberg, 2023 (10) | Cohort; no | Ghana | Inpatient | Mixed | 154 | 6.1 | 100 | 7.8 | NR | 1 | Spot | 0.27 (0.20, 0.35) | NR | Low, High |
| Alcantara 2012 (11) | CSS; no | Brazil | Outpatient | Mixed | 265 | 18.9 | NR | 16.0 | NR | 2 | 2 days | 0.06 (0.03, 0.09) | NR | Low, Unsure |
| Ali, 2021 (12) | Cohort; no | Pakistan | Outpatient | Tertiary | 2896 | 0.93 | NR | 11.3 | Self-exp | 1 | Spot | 0.0 (0.0, 0.0) | NR | Low, Unsure |
| Alvarez, 2015 (13) | Cohort; yes | Canada | Mixed | Secondary | 344 | 7.9 | NR | NR | Induced if unable to exp | 1-3 | >2 days | 0.0 (0.0, 0.01) | NR | Low, Low |
| Baghaei, 2011 (14) | CSS; yes | Iran | NR | Tertiary | 809 | 61.0 | 0 | 0 | NR | NR | NR | 0.11 (0.08, 0.13) | NR | Low, High |
| Baik, 2020 (15) | C-C; no | Uganda | Outpatient | Primary | 425 | 27.4 | 33.9 | 14.5 | NR | NR | NR | 0.04 (0.02, 0.06) | NR | Low, High |
| Balcells, 2012 (16) | CSS; yes | Chile | Mixed | Mixed | 162 | 7.5 | 100 | 11.8 | Self-exp | 2 | Spot | 0.19 (0.13, 0.25) | NR | Low, Low |
| Balcha, 2014 (17) | Cohort; yes | Ethiopia | Outpatient | Primary | 873 | 16.9 | 100 | 6.0 | Self-exp | 4 | 2 days | 0.07 (0.05, 0.09) | NR | Low, High |
| Basir, 2019 (18) | CSS; no | Pakistan | Outpatient | Primary | 153 | 22.8 | NR | 13.9 | Self-exp | NR | NR | 0.25 (0.18, 0.32) | NR | Low, High |
| Bassett, 2012 (19) | Cohort; no | South Africa | Outpatient | NR | 951 | 15.0 | 100 | 26.0 | Induced if unable to exp | 1 | Spot | 0.0 (0.0, 0.0) | NR | Low, Low |
| Belay, 2015 (20) | CSS; no | Ethiopia | NR | Mixed | 339 | 32.3 | 28.6 | NR | NR | 3 | 2 days | 0.04 (0.02, 0.07) | NR | Low, Low |
| Benjamin 2019 (21) | CSS; yes | Brazil | Outpatient | Tertiary | 205 | 24.6 | 100 | 23.1 | Induced if unable to exp | 2 | NR | 0.06 (0.03, 0.10) | NR | Low, Low |
| Berhanu 2018 (22) | CSS; yes | South Africa | Outpatient | Mixed | 299 | 24.5 | 62 | 18.1 | Self-exp | 4 | >2 days | 0.03 (0.01, 0.06) | NR | Low, Low |

| | | | | | | | | | | | | | | |
|---|---|---|---|---|---|---|---|---|---|---|---|---|---|---|
| Bigogo, 2018 (23) | CSS; no | Kenya | Outpatient | Mixed | 7500 | 5.4 | 14.6 | NR | Self-exp | 1 | Spot | 0.83 (0.82, 0.84) | NR | Low, High |
| Bjerrum, 2015 (24) | Cohort; yes | Ghana | Mixed | Tertiary | 495 | 11.7 | 100 | 6.0 | Self-exp | 2 | >2 days | 0.04 (0.03, 0.06) | NR | Low, Low |
| Bonnet, 2011 (25) | CSS; yes | Kenya | Outpatient | Primary | 509 | 23.9 | NR | 20.6 | Self-exp | 4 | >2 days | 0.02 (0.01, 0.04) | 3.7 | Low, Low |
| Boum, 2013 (26) | CSS; yes | Uganda | Outpatient | Tertiary | 891 | 23.7 | 58.6 | 2.6 | NR | 2 | 2 days | 0.25 (0.22, 0.28) | 0.3 | Low, Low |
| Boyles, 2018 (27) | Cohort; yes | South Africa | Inpatient | Secondary | 358 | 50.9 | 100 | NR | Induced if unable to exp | 3 | NR | 0.07 (0.05, 0.10) | NR | Low, Low |
| Boyles, 2020 (28) | Cohort; yes | South Africa | Outpatient | Primary | 217 | 36.0 | 100 | 17.0 | Induced if unable to exp | 2 | >2 days | 0.0 (0.0, 0.02) | NR | Low, High |
| Burhan, 2022 (29) | CSS; no | Indonesia | Outpatient | Tertiary | 490 | 69.8 | 5.2 | 42.0 | Induced if unable to exp | NR | NR | 0.04 (0.02, 0.06) | NR | Low, Low |
| Calder-wood, 2023 (30) | Cohort; yes | South Africa | Outpatient | Primary | 1097 | 27.0 | 42.0 | 41.0 | Induced if unable to exp | 2 | NR | 0.08 (0.07, 0.10) | NR | Low, High |
| Carriquiry, 2012 (31) | CSS; yes | Peru | NR | Tertiary | 152 | 34.4 | 100 | 25.0 | Self-exp | 2 | 2 days | 0.11 (0.06, 0.17) | NR | Low, Low |
| Cattaman-chi, 2011 (32) | CSS; yes | Uganda | Inpatient | Tertiary | 492 | 50.0 | 69.0 | NR | NR | 2 | 2 days | 0.03 (0.02, 0.05) | 21.4 | Low, Low |
| Chaisson, 2015 (33) | CSS; yes | Vietnam | NR | Tertiary | 332 | 28.5 | 0 | NR | NR | 2 | NR | 0.0 (0.0, 0.01) | NR | Low, High |
| Chawla, 2016 (34) | CSS; no | Malawi | Inpatient | Tertiary | 658 | 9.1 | 30.4 | NR | Self-exp | 1 | Spot | 0.53 (0.49, 0.57) | NR | Low, Low |
| Chew, 2016 (35) | CSS; yes | Singapore | Inpatient | Tertiary | 450 | 13.6 | NR | NR | Induced if unable to exp | 2 | Spot | 0.0 (0.0, 0.01) | NR | Low, High |
| Chilukutu, 2022 (36) | CSS; no | Zambia | Outpatient | Primary | 771 | 12.4 | 43.1 | 19.3 | NR | 2 | Spot | 0.01 (0.00, 0.02) | NR | Low, High |
| Churchyard 2015 (37) | RCT; no | South Africa | Outpatient | Primary | 4677 | 8.7 | 62 | 15.4 | NR | 1-2 | NR | 0.0 (0.0, 0.01) | NR | Low, High |
| Cowan, 2017 (38) | Cohort; no | United States of America | Inpatient | Tertiary | 329 | 6.3 | 23.9 | NR | Self-exp | 1-3 | 2 days | 0.03 (0.01, 0.06) | NR | Low, Low |
| Cox, 2014 (39) | RCT; no | South Africa | Outpatient | Primary | 1985 | 21.5 | 48.6 | 37.5 | NR | 2 | NR | 0.02 (0.01, 0.03) | NR | Low, Low |

| | | | | | | | | | | | | | | |
|---|---|---|---|---|---|---|---|---|---|---|---|---|---|---|
| Cuevas, 2011 (40) | CSS; yes | Ethiopia, Nepal, Nigeria, Yemen | NR | Mixed | 6627 | 24.1 | 8.8 | NR | Self-exp | 3 | 2 days | 0.04 (0.04, 0.05) | NR | Low, Low |
| Demelash, 2023 (41) | CSS; yes | Ethiopia | Outpatient | Tertiary | 180 | 8.8 | NR | NR | Self-exp | 2 | Spot | 0.0 (0.0, 0.02) | NR | Unsure High |
| Der, 2021 (42) | CSS; no | Ghana | Outpatient | Secondary | 236 | 3.2 | 10.8 | 3.8 | Self-exp | 1 | Spot | 0.20 (0.15, 0.26) | NR | Low, Low |
| Divala, 2023 (43) | RCT; no | Malawi | Outpatient | Primary | 1583 | 6.3 | 14.9 | 6.3 | NR | 2 | >2 days | 0.18 (0.16, 0.20) | NR | Low, Low |
| Drain, 2014 (44) | CSS; yes | South Africa | Outpatient | Mixed | 399 | 17.5 | 100 | 7.6 | Induced if unable to exp | 1 | Spot | 0.11 (0.08, 0.15) | NR | Low, Low |
| Dutschke, 2022 (45) | CSS; no | Guinea-Bissau | Outpatient | Tertiary | 390 | 12.6 | 100 | 6.6 | Self-exp | 1 | >2 days | 0.48 (0.43, 0.53) | NR | Low, Low |
| El-Helbawy 2020 (46) | Cohort; yes | Egypt | NR | NR | 452 | 24.7 | NR | 8.0 | Induced if unable to exp | 3 | 2 days | 0.0 (0.0, 0.01) | 88.4 | Low, High |
| Fan, 2014 (47) | Cohort; yes | China | Inpatient | Tertiary | 335 | 27.5 | 0 | 2.0 | Self-exp | 3 | NR | 0.24 (0.19, 0.29) | NR | Low, Low |
| Farr, 2019 (48) | CSS; no | Uganda | Outpatient | Primary | 5330 | 8.5 | 47.9 | 0 | NR | NR | NR | 0.44 (0.43, 0.45) | NR | Low, High |
| Feasey, 2013 (49) | Cohort; yes | Malawi | Inpatient | Tertiary | 104 | 43.0 | 100 | 0 | Self-exp | 3 | 2 days | 0.19 (0.12, 0.28) | NR | Low, Low |
| Gammo, 2013 (50) | CSS; yes | Libya | NR | Tertiary | 412 | 23.5 | NR | NR | Self-exp | 4 | 2 days | 0.0 (0.0, 0.01) | NR | Low, Low |
| Gebreegzia biher, 2017 (51) | CSS; no | Ethiopia | Outpatient | NR | 201 | 0.6 | 21.9 | NR | Self-exp | 3 | 2 days | 0.13 (0.09, 0.19) | NR | Low, Low |
| Gounder, 2011 (52) | CSS; no | South Africa | Outpatient | Mixed | 678 | 0.4 | 36.7 | 4.8 | Self-exp | 1 | Spot | 0.51 (0.47, 0.54) | NR | Low, Low |
| Grant, 2020 (53) | RCT; no | South Africa | Outpatient | Primary | 1507 | 6.8 | 100 | 9.5 | Self-exp | 1 | Spot | 0.36 (0.33, 0.38) | NR | Low, High |
| Gray, 2016 (54) | CSS; yes | India, Uganda, Peru | NR | Mixed | 1995 | 22.1 | 22.2 | NR | NR | 2 | NR | 0.03 (0.02, 0.04) | NR | Low, Low |
| Gupta-Wright 2018 (55) | RCT; yes | Malawi, South Africa | Inpatient | Tertiary | 2574 | 11.5 | 100 | 25.0 | Self-exp | 1 | Spot | 0.43 (0.41, 0.45) | NR | Low, Low |

| | | | | | | | | | | | | | | |
|---|---|---|---|---|---|---|---|---|---|---|---|---|---|---|
| Hanifa, 2012 (56) | Cohort; no | South Africa | Outpatient | Secondary | 381 | 17.7 | 100 | 28.0 | Self-exp | 2 | Spot | 0.03 (0.01, 0.05) | NR | Low, Low |
| Hanifa, 2018 (57) | Cohort; no | South Africa | Outpatient | NR | 367 | 7.0 | 100 | 24.0 | Self-exp | 1 | Spot | 0.36 (0.31, 0.41) | NR | Low, Low |
| Hanifa, 2019 (58) | Cohort; no | South Africa | Outpatient | NR | 103 | 14.0 | 100 | 10.7 | Induced if unable to exp | 1 | Spot | 0.03 (0.01, 0.08) | NR | Low, Low |
| Huerga, 2017 (59) | Cohort; yes | Kenya | Mixed | Tertiary | 474 | 56.7 | 100 | 24.7 | Induced if unable to exp | 2 | 2 days | 0.23 (0.19, 0.27) | NR | Low, Low |
| Huerga, 2019 (60) | Cohort; no | Malawi, Mozambique | Outpatient | Mixed | 456 | 24.9 | 100 | NR | Self-exp | 2 | 2 days | 0.14 (0.11, 0.18) | NR | Low, Low |
| Huerga, 2020 (61) | Cohort; yes | Malawi | Outpatient | Mixed | 485 | 14.1 | 100 | NR | Self-exp | 2 | 2 days | 0.08 (0.06, 0.11) | NR | Low, Low |
| Huerga, 2021 (62) | CSS; yes | Malawi | Inpatient | Secondary | 387 | 30.8 | 100 | NR | Self-exp | 2 | >2 days | 0.34 (0.30, 0.39) | NR | Low, Low |
| Huerga, 2023 (63) | CSS; yes | Uganda, Kenya, Mozambique, South Africa | Outpatient | Mixed | 1031 | 9.4 | 100 | NR | Induced if unable to exp | 2 | Spot | 0.10 (0.08, 0.12) | NR | Low, Low |
| Jones-Lopez 2014 (64) | CSS; yes | Uganda | Inpatient | Tertiary | 212 | 27.1 | 81.6 | 11.3 | Self-exp | 3 | >2 days | 0.0 (0.0, 0.02) | 44.0 | Low, Low |
| Kalema, 2012 (65) | CSS; no | Uganda | Inpatient | Tertiary | 245 | 47.2 | 80.0 | NR | Self-exp | 2 | Spot | 0.08 (0.05, 0.12) | 13.7 | Low, Low |
| Kancheya, 2014 (66) | Cohort; no | Zambia | Outpatient | Primary | 1422 | 1.5 | 17.0 | 2.9 | Self-exp | 2 | Spot | 0.11 (0.09, 0.12) | NR | Low, Low |
| Kasaro, 2020 (67) | CSS; yes | Zambia | Outpatient | Mixed | 1350 | 15.4 | 100 | NR | Self-exp | 3 | NR | 0.20 (0.18, 0.22) | NR | High, Low |
| Kempker, 2019 (68) | CSS; no | Georgia | Outpatient | Tertiary | 131 | 11.5 | 100 | 3.0 | Self-exp | 2 | 2 days | 0.21 (0.15, 0.29) | NR | Low, Low |
| Khan, 2020 (69) | CSS; yes | Pakistan | Outpatient | Tertiary | 2368 | 12.4 | 0.5 | 23.0 | Induced if unable to exp | 3 | Spot | 0.02 (0.02, 0.03) | NR | Low, Low |
| Kweza, 2018 (70) | CSS; no | South Africa | Outpatient | Primary | 1255 | 5.4 | 17.7 | 14.4 | Self-exp | 1 | Spot | 0.28 (0.25, 0.30) | NR | Low, Low |
| Lawn, 2010 (71) | Cohort; no | South Africa | Outpatient | Primary | 241 | 31.5 | 100 | 23.2 | Induced if unable to exp | 2 | NR | 0.0 (0.0, 0.02) | NR | Low, High |

| | | | | | | | | | | | | | | |
|---|---|---|---|---|---|---|---|---|---|---|---|---|---|---|
| Lawn, 2012 (72) | CSS; yes | South Africa | Outpatient | Primary | 602 | 17.3 | 100 | 26.5 | Induced if unable to exp | 2 | Spot | 0.10 (0.08, 0.13) | NR | Low, Low |
| Lawn, 2017 (73) | Cohort; yes | South Africa | Inpatient | Secondary | 427 | 32.6 | 100 | 46.1 | Induced if unable to exp | 2 | Spot | 0.63 (0.58, 0.68) | NR | Low, Low |
| Lessells, 2017 (74) | RCT; no | South Africa | Outpatient | Primary | 1281 | 12.9 | 92.5 | 39.0 | Self-exp | 2 | Spot | 0.04 (0.03, 0.05) | NR | Low, Low |
| Li, 2023 (75) | CSS; yes | China | Outpatient | Primary | 396 | 25.5 | NR | 16.4 | Induced if unable to exp | 3 | 2 days | 0.0 (0.0, 0.01) | NR | Low, High |
| Lodha, 2022 (76) | CSS; yes | India | Outpatient | Tertiary | 114 | 43.9 | 1.8 | NR | Induced if unable to exp | NR | NR | 0.11 (0.06, 0.18) | NR | Low, Low |
| Lora, 2015 (77) | CSS; yes | Bolivia | Mixed | Tertiary | 134 | 44.9 | 100 | NR | Induced if unable to exp | 3 | >2 days | 0.13 (0.08, 0.19) | NR | Low, Low |
| Mateyo, 2022 (78) | CSS; no | Zambia | Outpatient | Primary | 771 | 13.0 | 67.0 | 19.0 | NR | 1 | NR | 0.01 (0.0, 0.02) | NR | Low, Low |
| Mathebula, 2020 (79) | CSS; no | Botswana | Outpatient | NR | 1863 | 10.9 | 100 | 10.9 | Self-exp | 4 | 2 days | 0.53 (0.51, 0.55) | 29.3 | Low, Low |
| Mbu, 2018 (80) | CSS; no | Cameroon | Outpatient | Tertiary | 1149 | 13.9 | 100 | NR | NR | 2 | 2 days | 0.18 (0.16, 0.21) | NR | Low, Low |
| Meyer, 2017 (81) | CSS; no | Uganda | Inpatient | Tertiary | 3572 | 22.0 | 66.0 | 12.0 | Induced if unable to exp | 2 | Spot | 0.0 (0.0, 0.0) | 16.0 | Low, High |
| Miremba, 2012 (82) | CSS; yes | Uganda | Outpatient | Tertiary | 231 | 50.7 | 35.7 | 8.1 | Self-exp | 3 | 2 days | 0.01 (0.0, 0.03) | NR | Low, Low |
| Mtwangambate, 2014 (83) | Cohort; no | Tanzania | Mixed | Tertiary | 121 | 5.8 | 59. | 7.4 | Self-exp | 2 | 2 days | 0.72 (0.63, 0.80) | NR | Low, Low |
| Munseri, 2011 (84) | Cohort; no | Tanzania | Inpatient | Tertiary | 258 | 32.0 | 100 | 12.4 | Self-exp | 3 | 2 days | 0.14 (0.10, 0.19) | NR | Low, Low |
| Mupfumi, 2014 (85) | RCT; yes | Zimbabwe | Outpatient | Tertiary | 440 | 21.0 | 100 | NR | Induced if unable to exp | 2 | Spot | 0.04 (0.02, 0.06) | NR | Low, Low |
| Muyoyeta, 2021 (86) | CSS; yes | Zambia | Outpatient | Primary | 157 | 22.5 | 46.0 | 21.2 | Self-exp | 1 | Spot | 0.0 (0.0, 0.02) | NR | Low, High |
| Nabeta, 2017 (87) | CSS; yes | Peru, Vietnam | Outpatient | Mixed | 596 | 60.3 | NR | NR | NR | 2 | Spot | 0.04 (0.02, 0.06) | NR | Low, Low |
| Nakiyingi, 2014 (88) | CSS; yes | Uganda, South Africa | Mixed | Mixed | 1013 | 36.8 | 100 | 19.0 | Induced if unable to exp | 2 | Spot | 0.02 (0.01, 0.03) | NR | Low, Low |

| | | | | | | | | | | | | | | |
|---|---|---|---|---|---|---|---|---|---|---|---|---|---|---|
| Ngangue, 2022 (89) | CSS; yes | Cameroon | Outpatient | Secondary | 1030 | 27.0 | 37.0 | 14.0 | Self-exp | 2 | 2 days | 0.05 (0.04, 0.07) | NR | Low, Low |
| Nguyen, 2014 (90) | CSS; no | Vietnam | Inpatient | Tertiary | 94 | 23.4 | 75.0 | NR | Self-exp | NR | NR | 0.35 (0.26, 0.46) | NR | Low, Low |
| Pandey, 2019 (91) | CSS; yes | India | NR | Mixed | 290 | 30.0 | 0 | NR | NR | 2 | 2 days | 0.18 (0.14, 0.23) | NR | High, High |
| Pant, 2022 (92) | CSS; no | Nepal | NR | Tertiary | 104 | 9.6 | NR | NR | NR | NR | NR | 0.0 (0.0, 0.03) | NR | High, High |
| Penn-Nicholoson 2021 (93) | CSS; yes | Peru, India, Ethiopia, Papua New Guinea | Outpatient | Primary | 1904 | 24.0 | 5.3 | NR | Self-exp | 4 | 2 days | 0.05 (0.04, 0.06) | NR | Low, Low |
| Peter, 2012 (94) | CSS; yes | South Africa | Inpatient | Mixed | 281 | 48.0 | 100 | 35.0 | Self-exp | 2 | NR | 0.26 (0.21, 0.32) | NR | Low, Low |
| Peter, 2016 (95) | RCT; yes | South Africa, Tanzania, Zambia, Zimbabwe | Inpatient | Secondary | 2528 | 29.2 | 100 | 27.0 | Induced if unable to exp | 2 | NR | 0.07 (0.06, 0.08) | NR | Low, Low |
| Quinco, 2013 (96) | CSS; yes | Brazil | Mixed | Tertiary | 508 | 19.4 | 60.4 | 3.0 | NR | 2 | 2 days | 0.02 (0.01, 0.03) | NR | Low, Low |
| Rachow, 2022 (97) | Cohort yes | Romania | Outpatient | Tertiary | 139 | 42.4 | 0.7 | 29.5 | Self-exp | 2 | Spot | 0.0 (0.0, 0.03) | NR | Low, Low |
| Reddy, 2010 (98) | CSS; no | Peru | NR | NR | 471 | 6.2 | 100 | 14.3 | Self-exp | 4 | >2 days | 0.04 (0.02, 0.06) | NR | Low, Low |
| Reddy, 2017 (99) | CSS; yes | South Africa | Outpatient | Primary | 717 | 23.3 | NR | 32.6 | Self-exp | 2 | Spot | 0.01 (0.0, 0.02) | NR | Low, Low |
| Reeve, 2023 (100) | CSS; yes | South Africa | Outpatient | Primary | 897 | 12.0 | 100.0 | 14.0 | Induced if unable to exp | 3 | Spot | 0.02 (0.02, 0.04) | NR | Low, Low |
| Sander, 2019 (101) | CSS; no | Cameroon | Outpatient | Tertiary | 1255 | 3.6 | 7.0 | 2.7 | Self-exp | 2 | 2 days | 0.27 (0.24, 0.29) | NR | Low, High |
| Sani, 2020 (102) | CSS; no | Nigeria | Outpatient | Tertiary | 216 | 19.9 | 0 | NR | NR | 3 | >2 days | 0.0 (0.0, 0.02) | NR | Low, High |
| Santoso, 2017 (103) | CSS; yes | Indonesia | NR | Tertiary | 40 | 40.0 | 100 | NR | Induced if unable to exp | 1 | Spot | 0.0 (0.0, 0.08) | NR | Low, Low |
| Scott, 2011 (104) | Cohort; yes | South Africa | Outpatient | Primary | 319 | 37.6 | 70.0 | NR | Self-exp | 3 | >2 days | 0.03 (0.01, 0.05) | NR | Low, Low |

| | | | | | | | | | | | | | | |
|---|---|---|---|---|---|---|---|---|---|---|---|---|---|---|
| Seong, 2014 (105) | RCT; no | Republic of Korea | Outpatient | Tertiary | 38 | 71.1 | 0 | NR | Self-exp | 3 | >2 days | 0.08 (0.02, 0.20) | NR | Low, Low |
| Shah, 2020 (106) | CSS; yes | South Africa, Uganda, India, Peru | Outpatient | NR | 1086 | 32.0 | 47.0 | NR | Self-exp | 1 | Spot | 0.02 (0.01, 0.03) | NR | Low, Low |
| Shinu, 2013 (107) | CSS; yes | India | Outpatient | Tertiary | 1184 | 31.5 | 0 | NR | Self-exp | 2 | 2 days | 0.01 (0.00, 0.01) | 5.3 | Low, Low |
| Solari, 2019 (108) | CSS; no | Peru | Outpatient | Primary | 237 | 5.1 | 0.9 | 8.6 | Self-exp | 1 | Spot | 0.12 (0.08, 0.17) | NR | Low, Low |
| Songkhla, 2019 (109) | CSS; yes | Thailand | Mixed | Tertiary | 308 | 25.7 | 100 | NR | Self-exp | 1 | NR | 0.07 (0.05, 0.11) | NR | Low, Low |
| Spooner, 2022 (110) | CSS; yes | South Africa | Outpatient | Mixed | 783 | 12.0 | 100 | 15.0 | Induced if unable to exp | 2 | Spot | 0.03 (0.02, 0.04) | NR | Low, Low |
| Theron, 2011 (7) | CSS; yes | South Africa | Outpatient | Primary | 496 | 29.0 | 31.0 | 34.0 | NR | 2 | Spot | 0.38 (0.33, 0.42) | NR | Low, High |
| Vadwai, 2012 (111) | CSS; yes | India | NR | Tertiary | 468 | 64.7 | NR | NR | Self-exp | 1 | NR | 0.04 (0.02, 0.06) | NR | Low, High |
| vanHoving, 2020 (112) | CSS; no | South Africa | Outpatient | Secondary | 424 | 41.5 | 100 | NR | Induced if unable to exp | 2 | 2 days | 0.44 (0.39, 0.49) | NR | Low, Low |
| vanLettow, 2015(113) | Cohort; no | Malawi | Outpatient | Mixed | 348 | 15.0 | 55.0 | 21.0 | NR | NR | NR | 0.07 (0.05, 0.10) | NR | Low, High |
| Vijayageeth a, 2019 (114) | CSS; no | India | Outpatient | Tertiary | 77 | 1.3 | 0.1 | 0.6 | Self-exp | 1 | Spot | 0.91 (0.82, 0.96) | NR | Low, Low |
| Wake, 2022 (115) | CSS; yes | South Africa | Mixed | Tertiary | 181 | 17.0 | 100 | 12.4 | Induced if unable to exp | NR | NR | 0.50 (0.42, 0.57) | NR | High, High |
| Wang, 2016 (116) | CSS; yes | China | NR | NR | 270 | 62.4 | NR | NR | Self-exp | 1 | NR | 0.04 (0.02, 0.07) | NR | Unsure High |
| Zu, 2019 (117) | CSS; no | China | NR | Tertiary | 440 | 55.7 | NR | NR | Self-exp | 3 | 2 days | 0.00 (0.00, 0.01) | NR | Low, Low |
| Yeong, 2020 (118) | CSS; yes | Australia | Inpatient | Tertiary | 64 | 16.1 | 0 | 23.2 | Self-exp | 3 | 2 days | 0.08 (0.03, 0.17) | NR | Low, Low |

| | | | | | | | | | | | | | | |
|---|---|---|---|---|---|---|---|---|---|---|---|---|---|---|
| Yu, 2023 (119) | Cohort no | China | NR | Tertiary | 158 | 51.3 | 0 | 46.9 | Self-exp | NR | NR | 0.20 (0.14, 0.27) | NR | Low, Low |
| Zhao, 2022 (120) | CSS; yes | China | NR | Tertiary | 605 | 25.9 | NR | NR | NR | NR | NR | 0.47 (0.43, 0.51) | NR | Low, High |

**Legend**: (abbreviations)
Design: CSS=cross-sectional study; RCT=randomized controlled trial; DTA=diagnostic test accuracy (yes, no)
Clinical setting: inpatient, outpatient, mixed inpatient and outpatient, not reported (NR)
Healthcare level: Primary, Secondary, Tertiary, mixed levels, not reported (NR)
N pts: number of participants attempting sputum collection
Collection method: self-expectorated, induced if unable to self-expectorate, not reported (NR)
Number of samples attempted: 1-2, more than 2, not reported (NR)
Time of sample collection: spot, 1-2 days, 2 days, >2 days, not reported (NR)
Risk of Bias (RoB): Patient selection, Applicability

### Figure S1: Meta-analysis of sputum scarcity for collection of self-expectorated 1-2 spot sputum samples

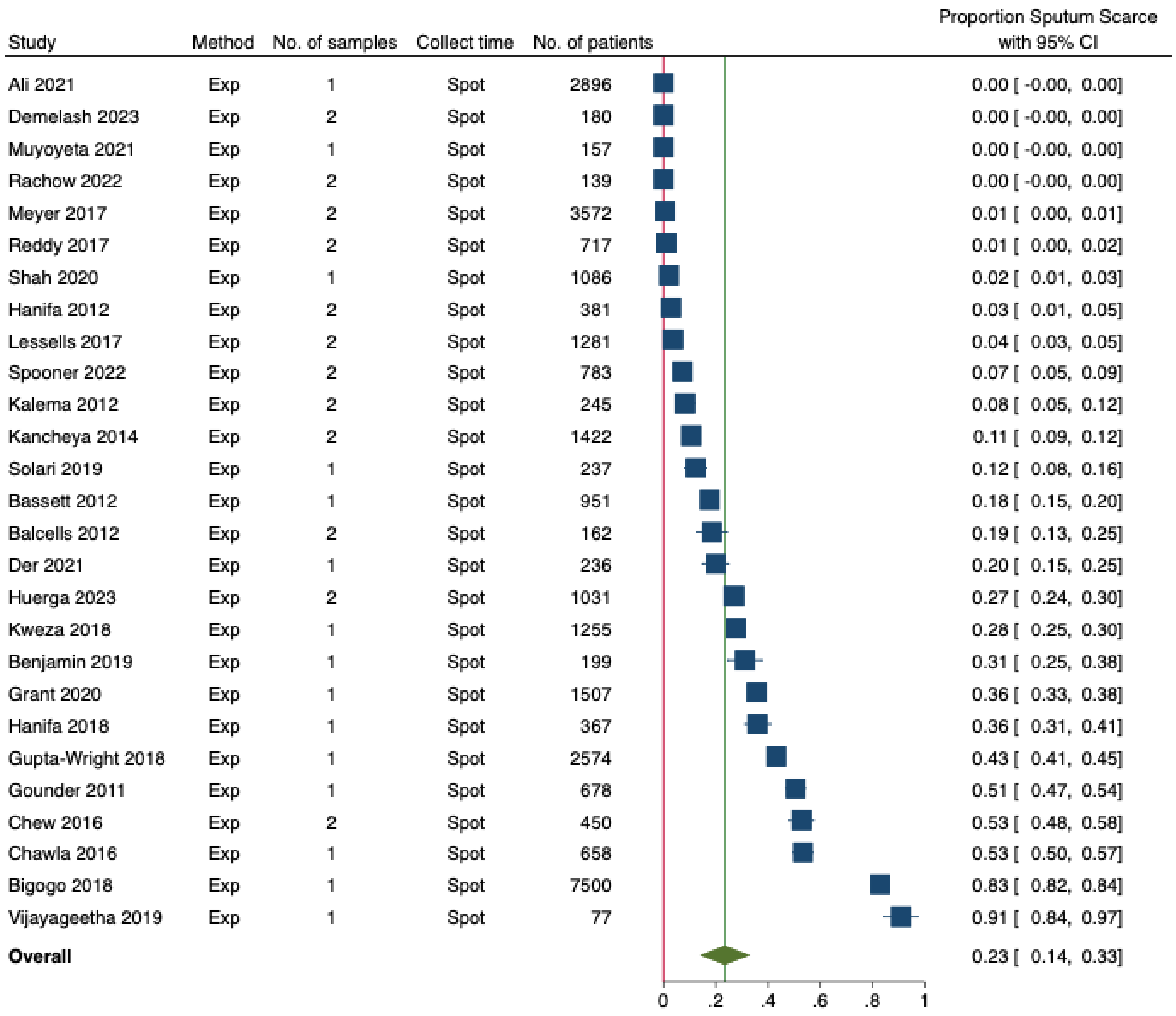


### Figure S2: Meta-analysis of sputum scarcity in people living with HIV for collection of 1-2 self-expectorated spot sputum samples

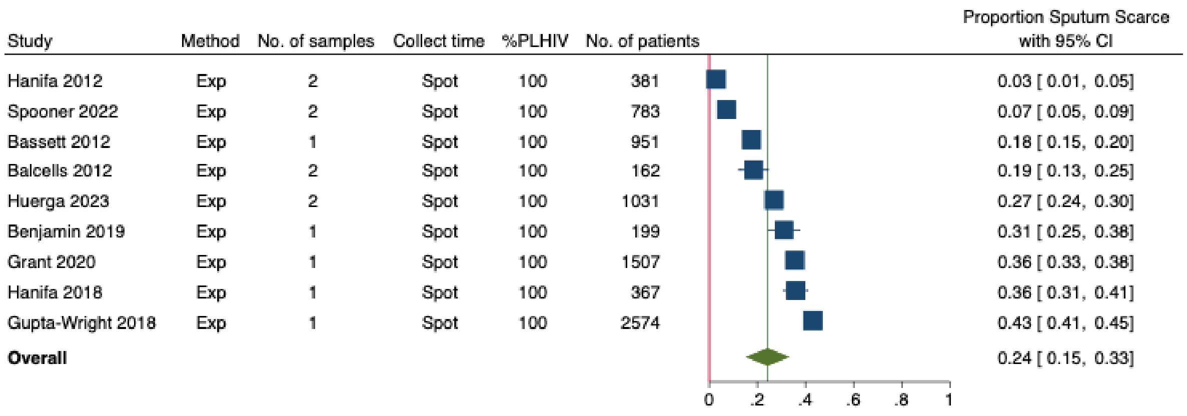

**Figure S3: Meta-analysis of sputum scarcity for studies with mixed HIV status for collection of 1-2 self-expectorated spot sputum samples**

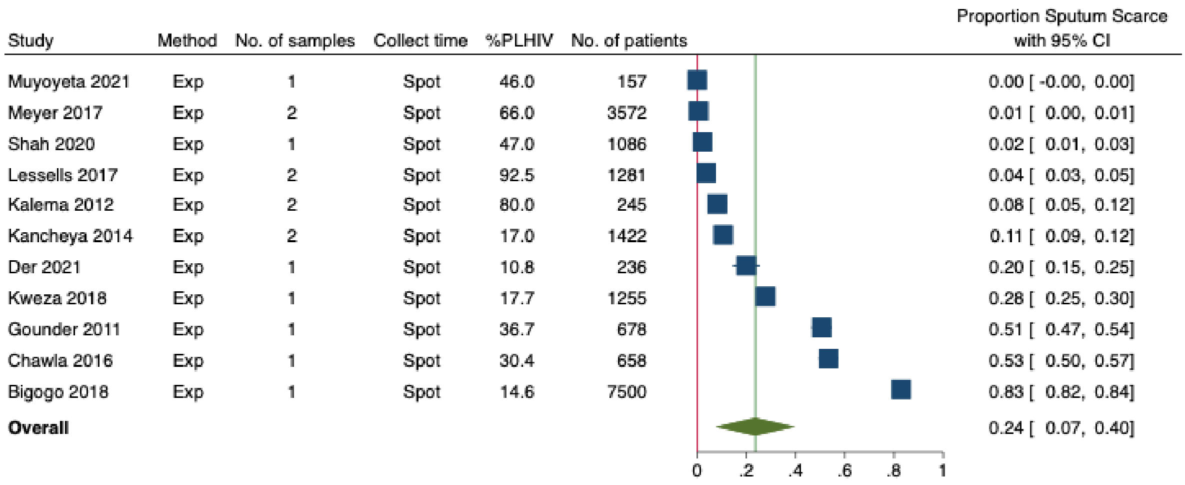


**Figure S4. Meta-analysis of sputum scarcity in studies enrolling PLHIV before initiation of anti-retroviral therapy for collection of 1-2 self-expectorated sputum samples**

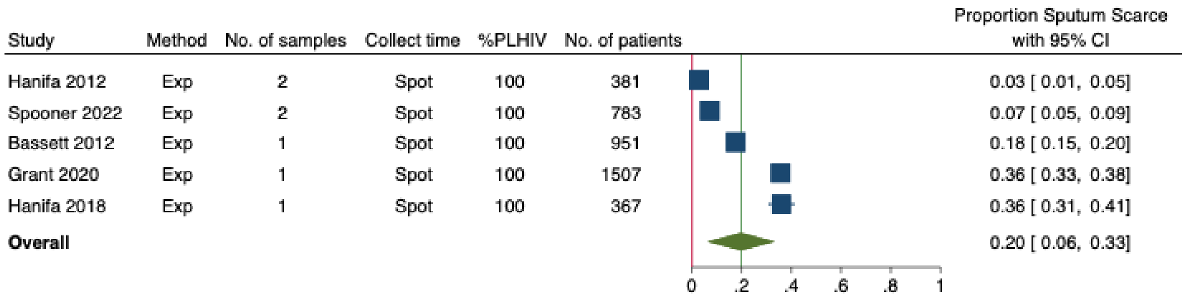


**Figure S5: Meta-analysis of sputum scarcity for people not living with HIV for collection of any self-expectorated sputum samples**

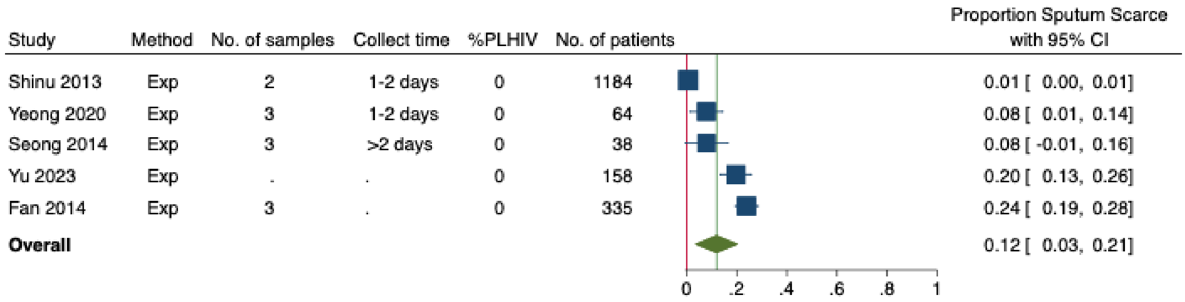

**Figure S6. Meta-analysis of sputum scarcity in people living with HIV with advanced disease and/or inpatient settings for collection of 1-2 self-expectorated sputum samples**

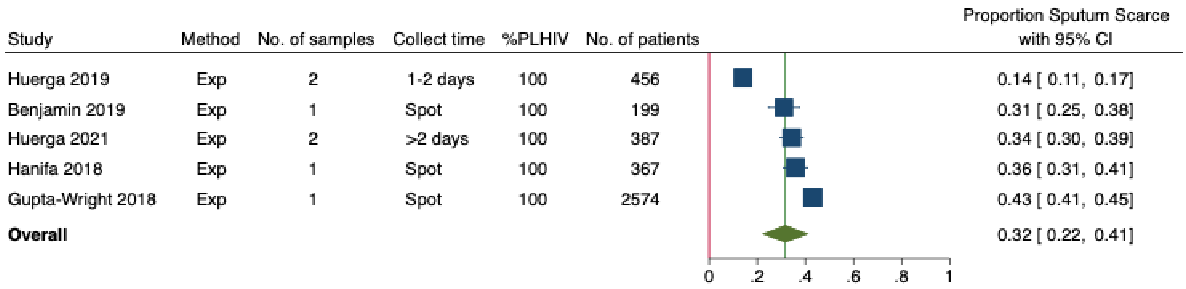


**Figure S7. Meta-analysis of sputum scarcity in outpatient settings for collection of self-expectorated 1-2 spot sputum samples**

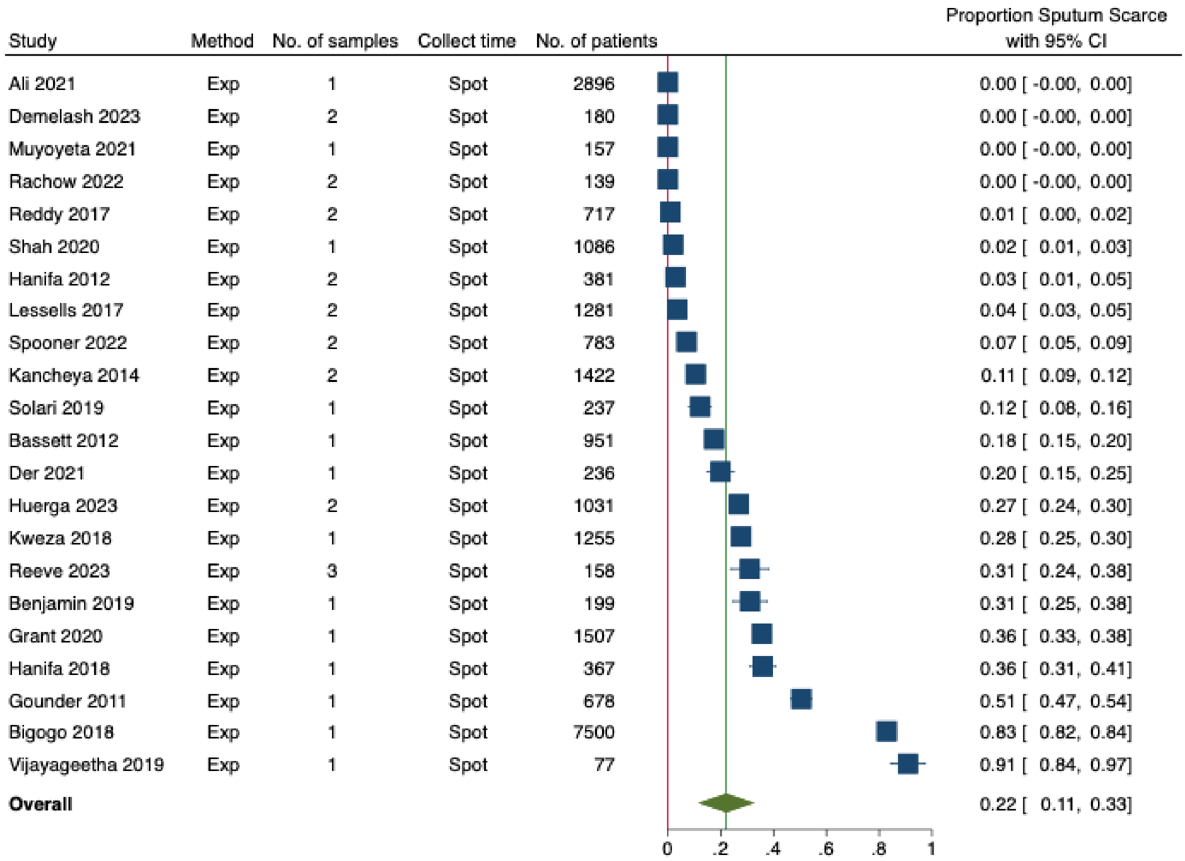

### Figure S8: Meta-analysis of sputum scarcity for collection of induced 1-2 spot samples

| Study | Method | No. of samples | Collect time | No. of patients | Proportion Sputum Scarce with 95% CI |
|---|---|---|---|---|---|
| Bassett 2012 | Ind | 1 | Spot | 167 | 0.00 [ -0.00, 0.00] |
| Meyer 2017 | Ind | 2 | Spot | 20 | 0.00 [ -0.00, 0.00] |
| Santoso 2017 | Ind | 1 | Spot | 40 | 0.00 [ -0.00, 0.00] |
| Nakiyingi 2014 | Ind | 2 | Spot | 1013 | 0.02 [ 0.01, 0.02] |
| Hanifa 2019 | Ind | 1 | Spot | 103 | 0.03 [ -0.00, 0.06] |
| Mupfumi 2014 | Ind | 2 | Spot | 440 | 0.04 [ 0.02, 0.05] |
| Chew 2016 | Ind | 2 | Spot | 238 | 0.07 [ 0.07, 0.07] |
| Benjamin 2019 | Ind | 1 | Spot | 62 | 0.10 [ -0.09, 0.29] |
| Lawn 2012 | Ind | 2 | Spot | 602 | 0.10 [ 0.01, 0.19] |
| Drain 2014 | Ind | 1 | Spot | 399 | 0.11 [ 0.08, 0.14] |
| Spooner 2022 | Ind | 2 | Spot | 56 | 0.18 [ 0.08, 0.28] |
| Lawn 2017 | Ind | 2 | Spot | 427 | 0.63 [ 0.59, 0.67] |
| **Overall** | | | | | 0.10 [ 0.00, 0.21] |

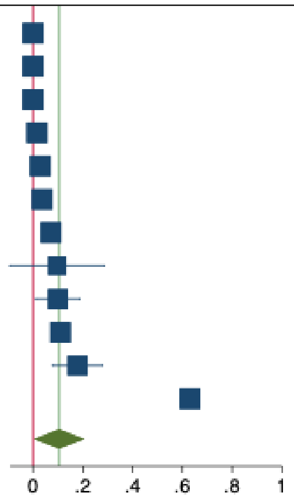


### Figure S9. Meta-analysis of sputum scarcity for collection of self-expectorated 1-2 spot samples from studies in high TB-burden countries

| Study | Country | Method | No. of samples | Collect time | No. of patients | Proportion Sputum Scarce with 95% CI |
|---|---|---|---|---|---|---|
| Ali 2021 | Pakistan | Exp | 1 | Spot | 2896 | 0.00 [ -0.00, 0.00] |
| Demelash 2023 | Ethiopia | Exp | 2 | Spot | 180 | 0.00 [ -0.00, 0.00] |
| Muyoyeta 2021 | Zambia | Exp | 1 | Spot | 157 | 0.00 [ -0.00, 0.00] |
| Meyer 2017 | Uganda | Exp | 2 | Spot | 3572 | 0.01 [ 0.00, 0.01] |
| Reddy 2017 | South Africa | Exp | 2 | Spot | 717 | 0.01 [ 0.00, 0.02] |
| Shah 2020 | South Africa | Exp | 1 | Spot | 1086 | 0.02 [ 0.01, 0.03] |
| Hanifa 2012 | South Africa | Exp | 2 | Spot | 381 | 0.03 [ 0.01, 0.05] |
| Lessells 2017 | South Africa | Exp | 2 | Spot | 1281 | 0.04 [ 0.03, 0.05] |
| Spooner 2022 | South Africa | Exp | 2 | Spot | 783 | 0.07 [ 0.05, 0.09] |
| Kalema 2012 | Uganda | Exp | 2 | Spot | 245 | 0.08 [ 0.05, 0.12] |
| Kancheya 2014 | Zambia | Exp | 2 | Spot | 1422 | 0.11 [ 0.09, 0.12] |
| Bassett 2012 | South Africa | Exp | 1 | Spot | 951 | 0.18 [ 0.15, 0.20] |
| Huerga 2023 | Uganda | Exp | 2 | Spot | 1031 | 0.27 [ 0.24, 0.30] |
| Kweza 2018 | South Africa | Exp | 1 | Spot | 1255 | 0.28 [ 0.25, 0.30] |
| Benjamin 2019 | Brazil | Exp | 1 | Spot | 199 | 0.31 [ 0.25, 0.38] |
| Grant 2020 | South Africa | Exp | 1 | Spot | 1507 | 0.36 [ 0.33, 0.38] |
| Hanifa 2018 | South Africa | Exp | 1 | Spot | 367 | 0.36 [ 0.31, 0.41] |
| Gounder 2011 | South Africa | Exp | 1 | Spot | 678 | 0.51 [ 0.47, 0.54] |
| Bigogo 2018 | Kenya | Exp | 1 | Spot | 7500 | 0.83 [ 0.82, 0.84] |
| Vijayageetha 2019 | India | Exp | 1 | Spot | 77 | 0.91 [ 0.84, 0.97] |
| **Overall** | | | | | | 0.22 [ 0.10, 0.33] |

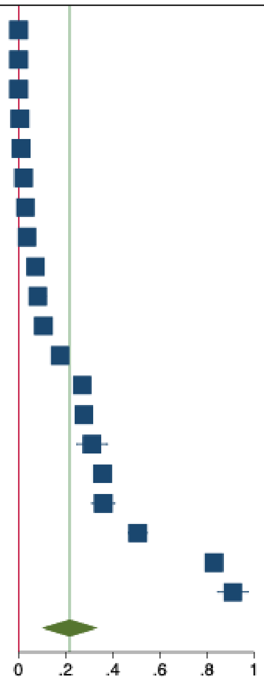

**Figure S10. Meta-analysis of sputum scarcity for collection of self-expectorated 1-2 spot samples from studies in not high TB-burden countries**

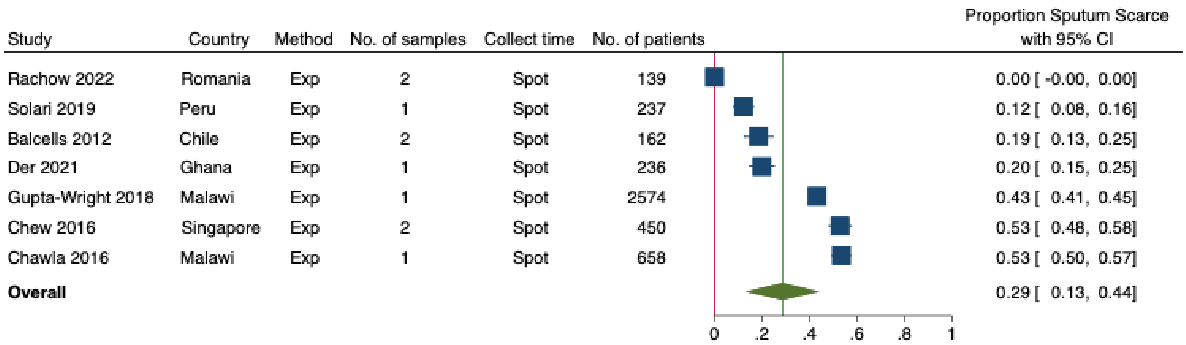


**Figure S11. Meta-analysis of sputum scarcity for collection of self-expectorated 1-2 spot sputum samples; sensitivity analysis removing 'zero scarcity' studies**

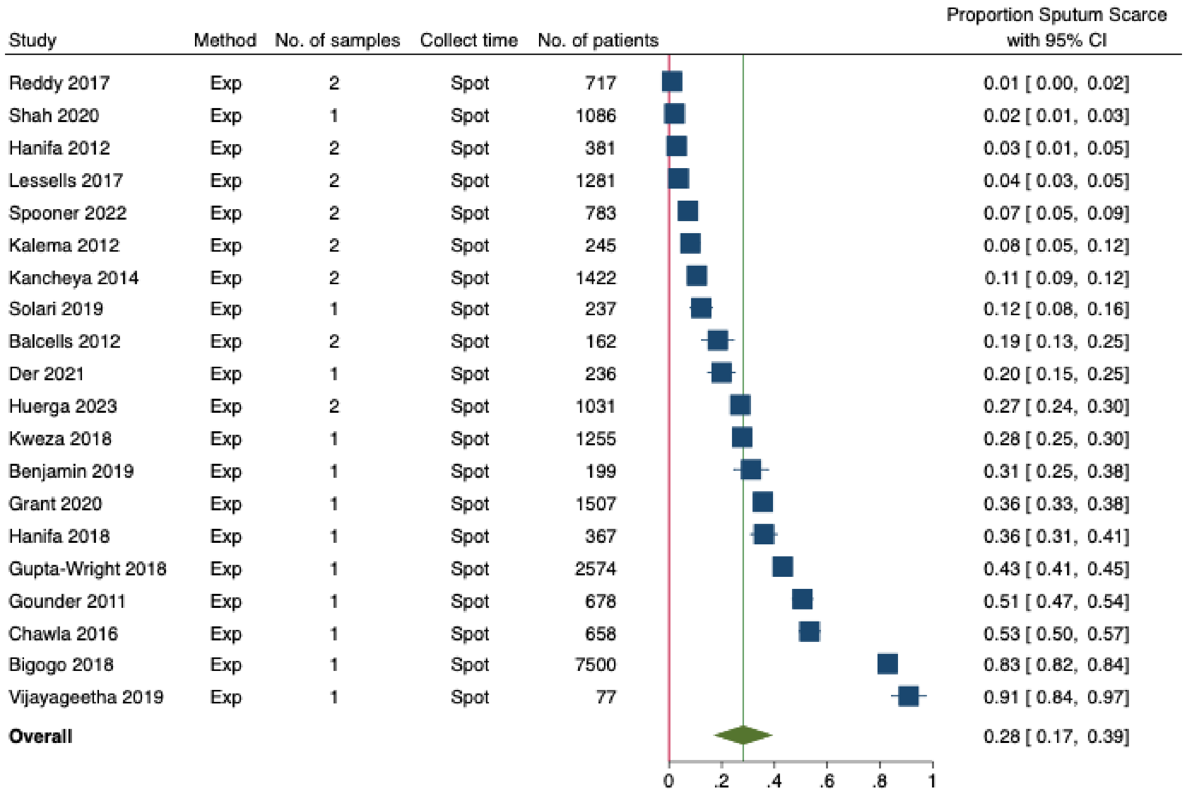

**Table S5. Sensitivity analyses**

| Sub-group | Number of studies | Proportion Sputum Scarce (95%CI) |
|---|---|---|
| All patients, Self-expectorated, Spot, 1-2 samples (original estimate) | 27 | 0.23 (0.14, 0.33) |
| All patients, Self-expectorated, 1-2 spot, removing studies with high Risk of Bias | 21 | 0.22 (0.12, 0.32) |
| All patients, Self-expectorated, 1-2 spot, removing studies reporting sputum collection from all ("zero studies") | 20 | 0.28 (0.17, 0.39) |
| Outpatients, Self-expectorated, 1-2 spot, removing studies reporting sputum collection from all ("zero studies") | 16 | 0.28 (0.14, 0.41) |
